# Statistical analysis plan for the Preventing InfusAte injuries Throughout a Child’s Hospitalisation (PATCH) trial: a type 1 hybrid randomised controlled trial

**DOI:** 10.64898/2026.11.15.26367580

**Authors:** Mari Takashima, Amanda J. Ullman, Toni Day, Robert S. Ware

## Abstract

**Background:** Up to 20% of hospitalised infants with a peripheral intravenous catheter (PIVC) have an infusate-related injury. The PATCH trial is testing whether an IV biosensor (ivWatch®) added to standard observation reduces infiltration and extravasation injuries compared with standard observation alone.

**Methods:** PATCH is a multicentre, type 1 hybrid effectiveness-implementation randomised controlled trial at three Queensland hospitals. Infants under 1 year (≥1000 g) with a PIVC expected to remain more than 24 hours for intermediate- or high-risk infusates, antimicrobials or continuous fluids are randomised 1:1, stratified by site (four strata). The target sample is 532.

**Analysis:** The primary outcome is any infiltration/extravasation injury (mild to severe harm, CCHMC Extravasation Harm Scale), confirmed by a blinded outcome assessment committee. The primary analysis follows intention to treat and uses logistic regression adjusted for randomisation stratum. The risk difference is estimated by standardisation, with the odds ratio also reported. Secondary outcomes include injury severity, time to PIVC failure, dwell time, replacement PIVC, quality of life, skin injuries and serious adverse events. This plan was finalised on 10/11/2026, during recruitment and before database lock or any unblinded analysis.

**Trial registration:** ACTRN12623000561684.

## Background

## 1. Administrative information

### 1.1 Purpose

This multi-site, superiority Type-1 hybrid randomised controlled trial (RCT) will test the effectiveness and explore the value and implementation contexts of an intravenous (IV) biosensor (ivWatch®, ivWatch LLC, Newport News, Virginia, USA), compared to standard care, to detect extravasations and prevent extravasation injury. The purpose of this document is to minimise bias and ensure transparency and internal validity for the findings of the trial by defining and making publicly available the analysis approach prior to reviewing or analysing any trial data. This statistical analysis plan (SAP) will inform the analysis and reporting of the effectiveness findings of the trial. It provides a detailed description of the primary and secondary trial effectiveness outcomes and the methods for statistical comparison.

### 1.2 Study identifiers

- Protocol: version 3.1, 28/04/2026
- ANZCTR registration number: ACTRN12623000561684

## 2. Study synopsis

This study is a multicentre, Type 1 hybrid effectiveness-implementation, superiority, parallel group, randomised controlled trial with a 1:1 randomisation to two arms: IV biosensor plus standard observation (intervention) or standard observation alone (control).

### 2.1 Study objectives

#### 2.1.1 Primary objective

The primary objective of this study is to compare the effectiveness of a novel IV biosensor + standard observation vs standard observation alone in reducing extravasation injuries in hospitalised infants.

#### 2.1.2 Secondary objectives

- To identify clinically effective and cost-effective methods to reduce extravasation injuries.
- To compare two observation methods [(i) novel IV biosensor + standard observation and (ii) standard observation alone].

#### 2.1.3 Primary hypothesis

- Hospitalised infants with IVs who have their site monitored by the IV biosensor + observation, in comparison to observation alone, will experience fewer extravasation injuries.

#### 2.1.4 Secondary hypothesis

- Hospitalised infants with IVs who have their site monitored by the IV biosensor + observation, in comparison to observation alone, will experience reduced extravasation severity, volume and treatment sequelae;
- The use of an IV biosensor for hospitalised infants will not be associated with an increase in replacement IVs.
- The IV biosensor will have lower healthcare costs at a micro-(patient) and macro-(health service) level compared to standard care.

#### 2.1.5 Inclusion criteria

- Neonates (≥1000 g, where the IV biosensor is physically able to be fitted to the limb) and infants (<1 year)
- Have an existing or new peripheral IV (including long and traditional peripheral IV) inserted in the distal half of a limb.
- Peripheral IV anticipated to be in situ for >24 hours.
- Planned administration via the study peripheral IV of either:

- intermediate to high risk infusates [1], as per the CHQ policy (see Figure 1). This includes intermediate (e.g., phenytoin, vancomycin, midazolam, morphine) and high-risk drugs (e.g., vasopressors, calcium, acyclovir, mannitol), or
- antimicrobials [2], or
- continuous fluids (including blood products) for 4 hours or more.

#### 2.1.6 Exclusion criteria

Participants who meet any of these criteria are not eligible for enrolment as trial participants:

- Other types of vascular access devices, including midline catheters, peripheral arterial catheters and central venous catheters
- Pre-existing skin condition affecting study peripheral IV site, e.g., i) atopic dermatitis or extensive erythema toxicum, ii) infectious diseases such as Staphylococcal scalded skin syndrome; iii) burn (e.g., heat, decontamination); iv) inherited conditions such as epidermolysis bullosa, ichthyosis and aplasia cutis congenita
- Previously enrolled in this study and randomised to one of the treatment conditions
- Currently receiving end-of-life, redirection of care, and palliative care.

**Figure 1.**
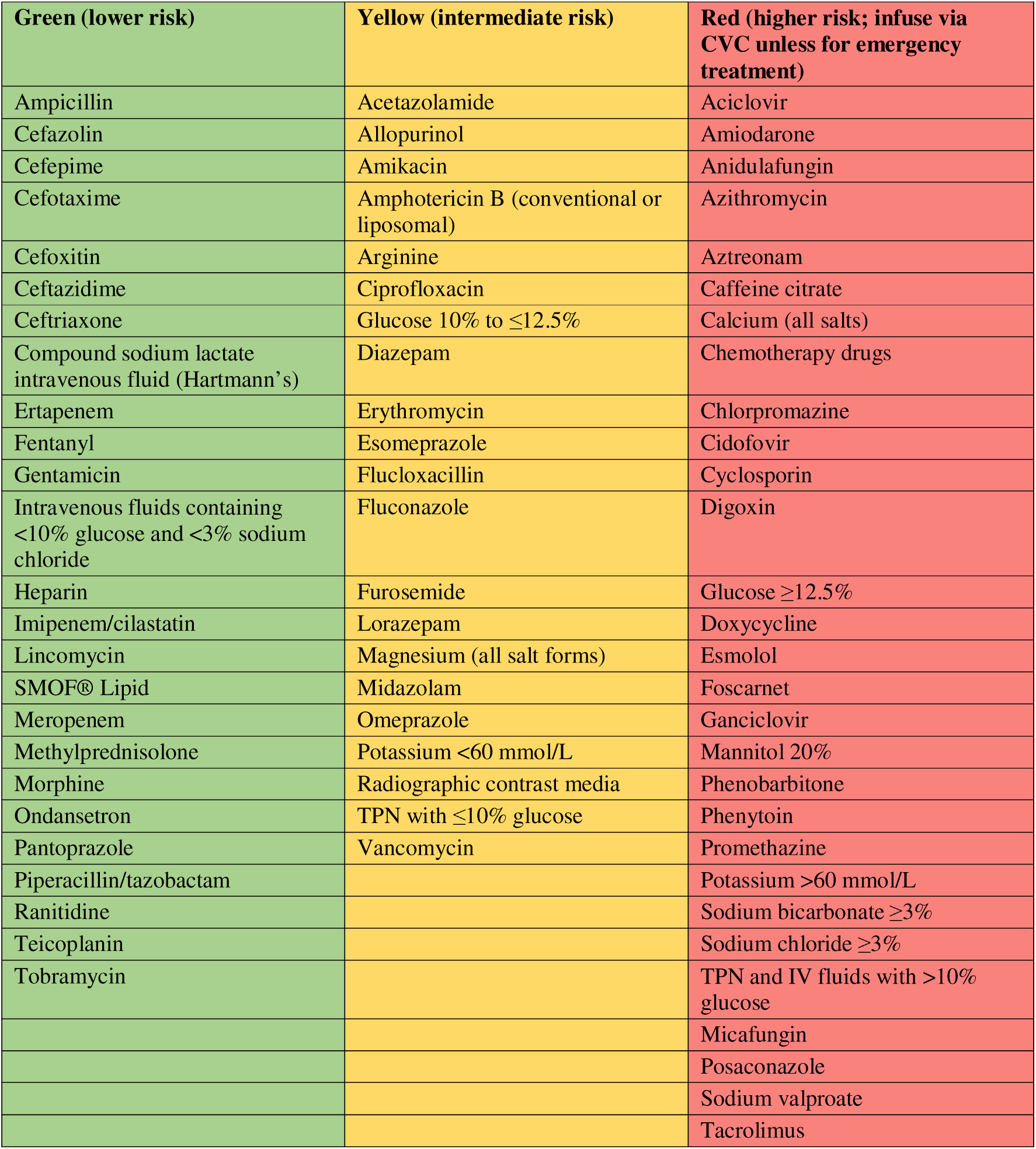
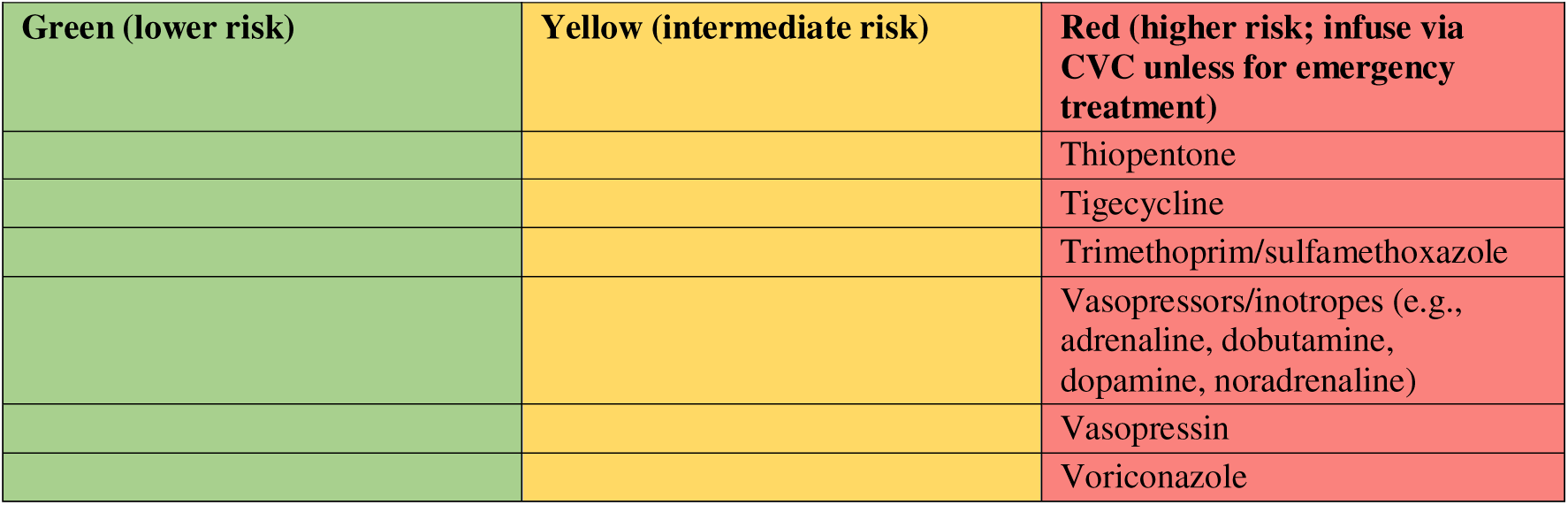
Infusate risk of phlebitis or local tissue injury due to infiltration/extravasation (CHQ-PROC-60579) The list is not exhaustive and includes commonly used medicines only; medicines not listed are checked with Pharmacy. CVC, central venous catheter; TPN, total parenteral nutrition.

### 2.2 Outcomes

#### 2.2.1 Primary outcome per protocol

The primary outcome is any extravasation, measured using the 4-point Cincinnati Children’s Hospital Medical Centre (CCHMC) Extravasation Harm Scale and then categorised as no harm vs any harm (that is, a composite of mild, moderate or severe harm) [1]. This scale combines physical (i.e., discolouration, swelling, redness, blisters, function, and arterial compromise), infusate properties (pH, osmolality, biological activity and volume) and injury treatment (scarring and surgical management) characteristics. The primary outcome is the Outcome Assessment Committee’s adjudicated rating; participants without a suspected injury referred to the Committee are classified as no harm.

#### 2.2.2 Secondary outcomes per protocol

These are the complications that occur during dwell, or at end of study completion, without failure, including:

1. Infiltration/extravasation severity: using the 4-point CCHMC Extravasation Harm Scale (no harm vs mild vs moderate vs severe).
2. Infiltration/extravasation volume: measured by the surface area to limb length ratio and photographic imaging, including comparison to the non-affected limb.
3. Infiltration/extravasation injury pH and colour changes: measured by the MGC tool and non-invasive monitors (pH meter).
4. Infiltration/extravasation treatment: measured by count of the number and type of dressings, skin grafting and/or scar management, and duration of treatment including scar management (censored at 6 months).
5. Requirement for replacement PIVC: yes/no, at PIVC removal.
6. Time to PIVC failure: hours from insertion to removal for PIVC dysfunction.
7. PIVC dwell time: hours from insertion to removal for any reason.
8. Near infrared/heat-related skin injuries: at the PIVC site (IV biosensor group only).
9. Mechanical adhesive-related and/or pressure injuries: surrounding the PIVC site.
10. Overall quality of life: PedsQL™ Infant Scales or IQI total score at recruitment and PIVC removal.
11. Serious adverse events (safety outcome): in-trial mortality, severe extravasation or device-related injury.

An exploratory analysis, not prespecified in the protocol, will compare outcomes 1 and 2 between participants with and without a red alarm in the IV biosensor group with an extravasation injury (Section 3.4.11).

### 2.3 Interventions

- Standard treatment: Standard clinical observation alone
- Intervention treatment: IV biosensor (sensor applied within 2.5 cm of the insertion site (4.5 cm for long PIVCs)) + standard clinical observation

Hereafter, these are referred to as the IV biosensor group and the standard care group.

This study will take place in two tertiary-referral hospitals (Queensland Children’s Hospital and Royal Brisbane and Women’s Hospital, Brisbane) and one secondary-referral hospital (Sunshine Coast University Hospital, Sunshine Coast).

### 2.4 Randomisation

Randomisation (patient level) will be a 1:1 ratio between groups, with randomly varied block sizes (6 to 8) and stratification by site, with SCUH stratified by unit (Neonatal Unit; Children’s and Adolescent Unit), giving four strata.

### 2.5 Data collection

Data will be directly input by the Research Nurse into a purpose-built case report form (CRF) in REDCap (Research Electronic Data Capture, Vanderbilt), hosted by the University of Queensland on password-protected devices. The Research Nurses will follow participants daily until PIVC removal. Data will be kept password-protected. The schedule of data collection is shown in Figure 2.

- At enrolment, Research Nurses will collect consent forms and data on patient characteristics (age, primary diagnosis, weight, skin colour, first or subsequent PIVC), provider characteristics (inserter workforce, insertion approach, planned device utility), PIVC characteristics (placement, location, catheter type, securement), and quality of life (PedsQL™ Infant Scales or IQI).
- Daily, Research Nurses will visit participants to assess for primary and secondary outcomes, current PIVC use, and any clinical, device or treatment factors changed since recruitment.
- Within 24 hours of PIVC removal, Research Nurses will collect data on PIVC dwell, reason for removal, PIVC dysfunction, need for replacement PIVC, near infrared skin injury, monitor alarms, mechanical adhesive related skin injury or pressure injury, serious adverse events including mortality, and quality of life. If an infiltration/extravasation injury is identified, the PIVC site will be assessed and documented.
- If an infiltration/extravasation injury occurs, the patient will be followed every 24 hours, collecting data on injury characteristics, volume, treatment and sequelae, until the injury stabilises.
- For participants with a confirmed extravasation injury only, additional follow-up questionnaires (quality of life and modified Brisbane Burns Scar Impact Profile) will be collected at 1, 3, and 6 months post-PIVC removal.

**Figure 2.**
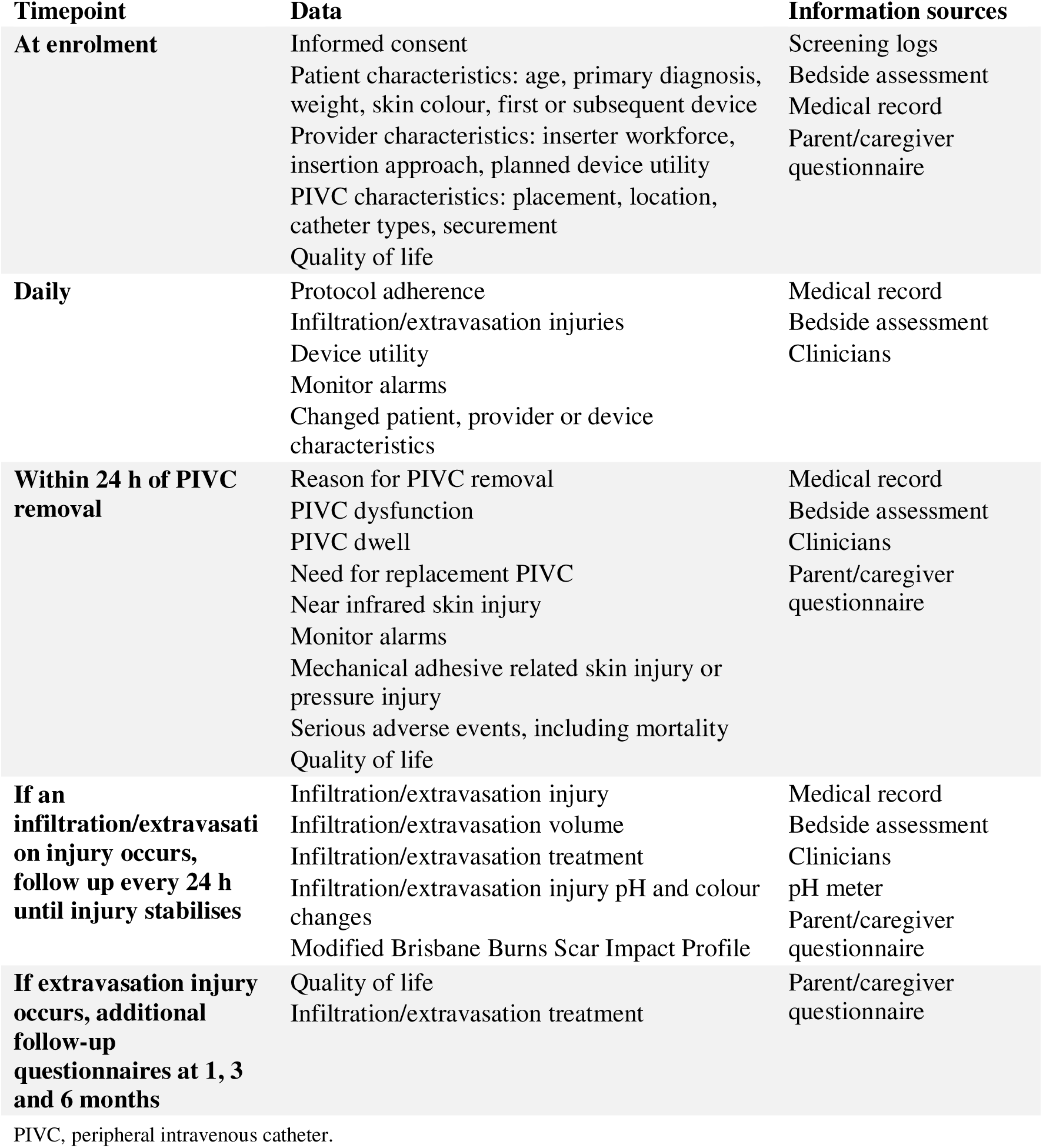
Schedule of data collection (adapted from the published PATCH protocol [**4**]) PIVC, peripheral intravenous catheter.

**Figure 3.**
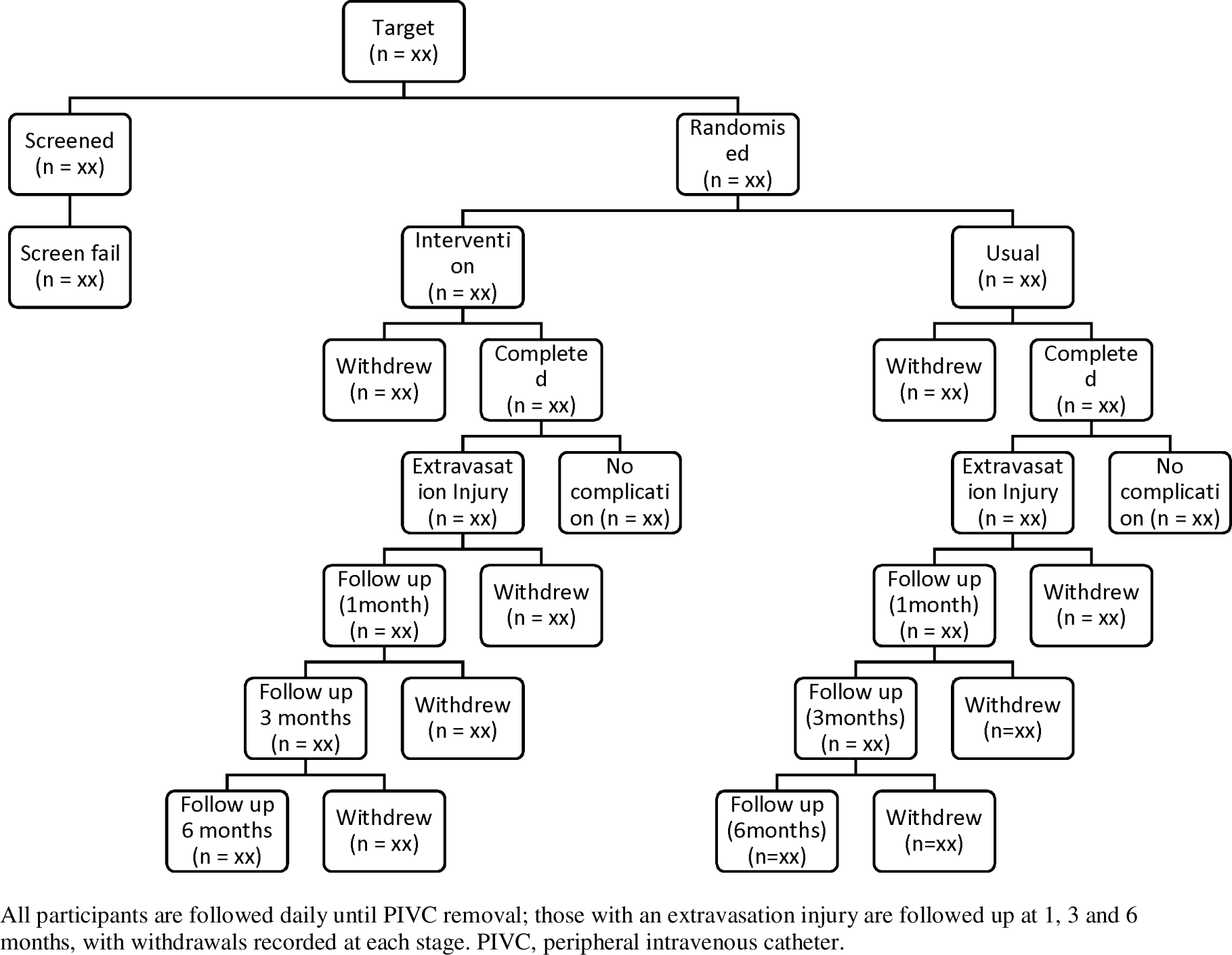
Participant follow-up. All participants are followed daily until PIVC removal; those with an extravasation injury are followed up at 1, 3 and 6 months, with withdrawals recorded at each stage. PIVC, peripheral intravenous catheter.

#### Outcome assessment process

On the day of recruitment, a high resolution clinical photograph of the PIVC site and comparator limb will be collected with study ID, date, time and a metric reference point using the MGC tool [3]. This baseline image will serve as a comparator for subsequent assessments, particularly in the event of potential infiltration/extravasation injury, PIVC dysfunction, or an IV biosensor Red Alert (IV biosensor arm only). If an injury is suspected (i.e., PIVC dysfunction, accidental device removal, catheter occlusion/blockage, inflammation at the insertion site, phlebitis, suspected extravasation, Red Alert), clinical details, additional images (baseline, and potential extravasation), clinical context (including PIVC function and signs of arterial compromise) and infusate properties (pH, osmolality, biological activity, and volume) will be reviewed by the blinded Outcome Assessment Committee (OAC) for verification. The OAC will determine injury status based on detailed definitions, and outcome attribution through group consensus and will assess whether long term follow up is required for the participant.

### 2.6 Sample size

Sample size calculations are based on binary expression of extravasation (no harm vs any (mild, moderate, severe) harm) [1]. Local and international data report approximately 30-42% of IVs with intermediate to high-risk infusates result in extravasation injuries in hospitalised infants under conditions similar to our ‘standard care’ scenario [5–8]. We conservatively estimate 20% of standard care alone participants will develop any extravasation injury (either mild, moderate, severe). Based on international observational [9,10] and our pilot RCT [11] data we expect 10% of IV biosensor will develop an extravasation injury. That is, we expect the IV biosensor to have a 50% relative risk reduction in extravasation, compared to standard observation alone. With α=0.05 and power=90%, we are required to record outcome data on 266 participants in each group to detect a between-group difference of this size or greater (total sample=532). Previous trials demonstrate it is feasible to recruit this number over two years across the three study sites.

### 2.7 Protocol violation

A protocol violation is defined as the non-randomised use or missed use of the IV biosensor.

## 3. Statistical analysis

### 3.1 General principles

All between-group regression models will include randomisation stratum (four levels) as a covariate, reflecting stratified randomisation. If a stratum has too few events for a model to converge, the two SCUH strata will be combined. Odds ratios and hazard ratios are therefore conditional on stratum.

#### 3.1.1 Intention to treat analysis

Analyses will follow intention to treat, with the PIVC as the unit of analysis (one PIVC per child). All randomised participants will be analysed in the group to which they were randomised, using all available data. The primary outcome will be analysed as complete case. As the primary outcome is assessed daily throughout dwell, missing primary outcome data are expected to be minimal, limited to participants who withdraw consent before any outcome assessment. Participants transferred before PIVC removal will be included, with the outcome assessed up to transfer (Figure 4). Safety outcomes will be reported for all randomised participants.

**Figure 4.**
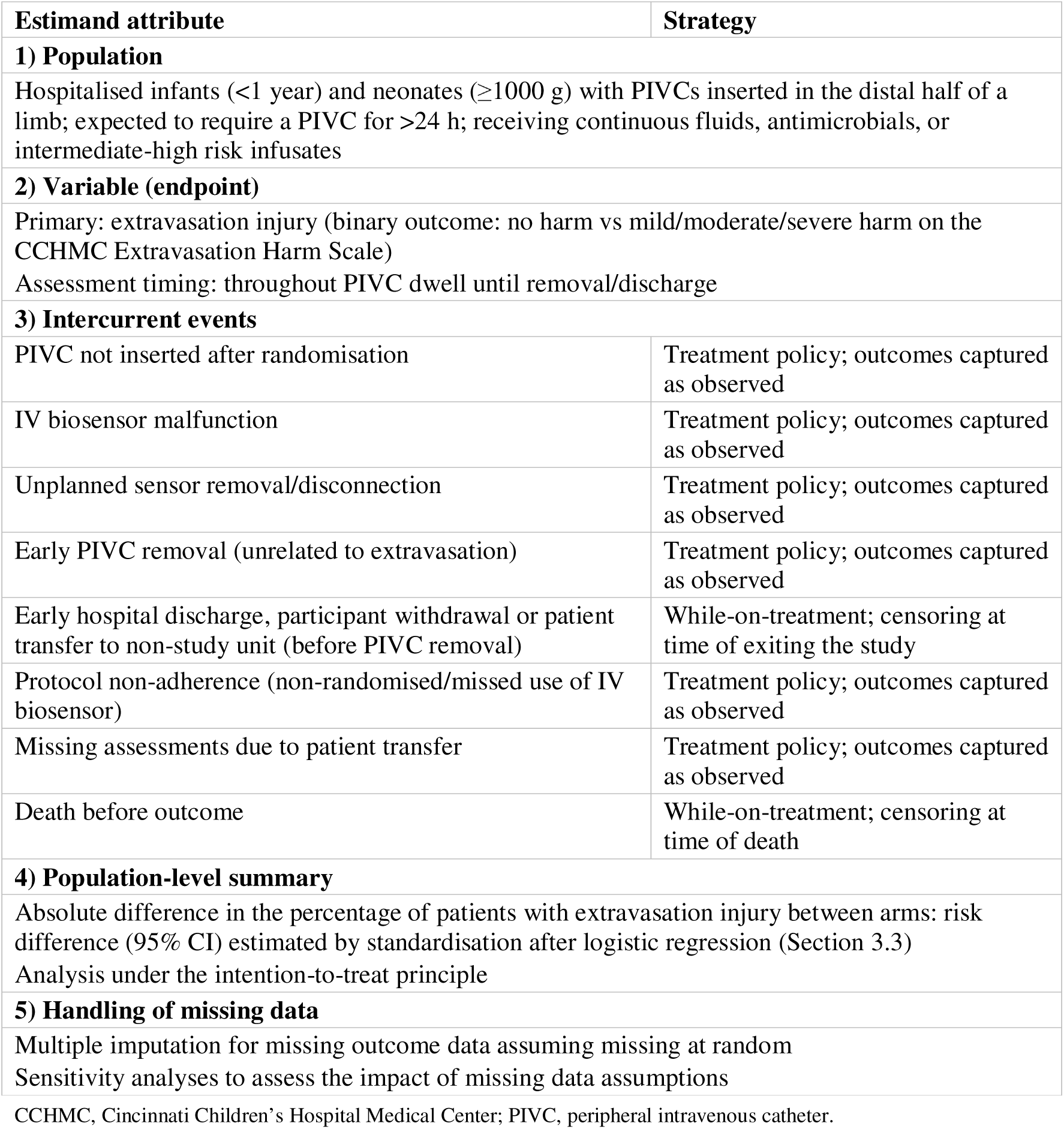
Estimand framework for the primary outcome (adapted from the published PATCH protocol [**4**]) CCHMC, Cincinnati Children’s Hospital Medical Center; PIVC, peripheral intravenous catheter.

Per protocol analysis will be performed to test the sensitivity of the results to noncompliance. This analysis will include participants who received their allocated intervention, excluding those with a protocol violation (Section 2.7).

The number of missing observations for each primary and secondary outcome will be reported by study group. If the proportion of missing observations is non-trivial (>5%), consideration will be given to the possible introduction of compliance and attrition bias. The cause of any missing data will be assessed.

#### 3.1.2 Stratified Analysis

For the primary outcome, between-group comparisons will be stratified by site to investigate the level of variability across sites. We will test the heterogeneity between sites by running a model with treatment group and site as main effects, and a group-by-site interaction term, and will report the level of interaction using the likelihood ratio test.

#### 3.1.3 Presentation of results

Continuous data will be summarised descriptively using either mean and standard deviation (SD) or median and interquartile range (IQR), depending on the distribution of the variable of interest. Categorical data will be presented as frequencies and percentages.

#### 3.1.4 Level of significance

A two-sided significance level of 0.05 will be used for the primary outcome. Secondary outcomes are exploratory; they will be reported with 95% CIs without adjustment for multiplicity.

#### 3.1.5 Data imputation

For any outcome with >5% missing data, a sensitivity analysis will use multiple imputation by chained equations (20 imputed datasets), performed separately by randomised group, assuming data are missing at random, with imputation models including treatment group, randomisation stratum, the outcome, age, weight and prematurity.

#### 3.1.6 Statistical software

All estimates will be derived using Stata (StataCorp, College Station, Texas, USA); version to be confirmed at time of analysis.

### 3.2 Evaluation of demographic and baseline characteristics

Demographic and baseline characteristics will be reported descriptively by allocated study group. Variable definitions are provided in Appendix 2 (data dictionary).

### 3.3 Planned analysis of the primary outcome

The primary outcome of any extravasation injury (no harm vs any harm on the CCHMC Extravasation Harm Scale) will be compared between groups using logistic regression, with treatment group (IV biosensor/standard care) as the main effect and randomisation stratum as a covariate. The effect estimate will be the absolute risk difference with 95% confidence interval and P-value, estimated by standardisation over the stratum distribution of all randomised participants (Stata margins) [12]. The odds ratio (95% CI), conditional on stratum, will also be reported from the same model.

### 3.4 Planned analysis of secondary outcomes

#### 3.4.1 Secondary outcome: Infiltration/extravasation severity

Infiltration/extravasation severity will be assessed using the 4-point CCHMC Extravasation Harm Scale, with each category (no harm, mild, moderate, or severe) compared. The distribution of severity categories will be reported by group as frequencies and percentages. An ordered logistic regression model with treatment group as the main effect and randomisation stratum as a covariate will be used to compare severity across groups, with results presented as odds ratios (95% CI).

#### 3.4.2 Secondary outcome: Infiltration/extravasation volume

Infiltration/extravasation volume, measured by the surface area to limb length ratio and photographic imaging (including comparison to the non-affected limb), will be summarised descriptively (median and IQR) by group for participants with an injury.

#### 3.4.3 Secondary outcome: Infiltration/extravasation treatment

Measured by count of the number and type of dressings, skin grafting and/or scar management, and duration of treatment, including scar management (censored at 6 months). As treatment is measured only in participants with an injury, the number and percentage of treatments will be reported descriptively by group, without between-group comparison. Treatment data are recorded on the stabilisation and follow-up forms.

#### 3.4.4 Secondary outcome: Infiltration/extravasation injury pH and colour changes

Measured by non-invasive monitors (pH Meter) and metric graduated colour (MGC) tool. As these are measured only in participants with an injury, they will be summarised descriptively by group (pH: median and IQR; colour: number and percentage).

#### 3.4.5 Secondary outcome: Requirement for replacement PIVC

Requirement for replacement PIVC (yes, including unsuccessful attempts, vs no) will be reported as number and percentage per group for all participants and will be analysed using logistic regression with study group as the main effect. The results will be presented as an odds ratio (95% CI).

#### 3.4.6 Secondary outcome: Time to PIVC failure and dwell time

Time to PIVC failure (removal for any PIVC dysfunction, including infiltration/extravasation) will be compared using Cox proportional hazards regression, with PIVCs removed at completion of therapy or for other reasons censored at removal. Time will be measured from PIVC insertion, with participants entering the risk set at randomisation (delayed entry), as most PIVCs were inserted before enrolment. Failure is recorded at the date and time of PIVC removal for dysfunction (infiltration/extravasation, other dysfunction, or a biosensor red alert); red alerts that did not lead to removal are not counted as failure. Results will be presented as hazard ratios (95% CI) and Kaplan-Meier curves. Dwell time (hours) will be summarised as median (IQR) by group and compared using quantile regression (median difference, 95% CI).

#### 3.4.7 Secondary outcome: Near infrared/heat-related skin injuries, at the PIVC site. The number and percentage of devices that experience near infrared/heat-related skin

injuries at the PIVC site will be reported descriptively (number and percentage) in the IV biosensor group only, as this injury can occur only with the sensor in place.

#### 3.4.8 Secondary outcome: Mechanical adhesive-related and/or pressure injuries, surrounding the PIVC site. The number and percentage of devices experiencing mechanical adhesive-related

skin injuries and pressure injuries surrounding the PIVC site will be calculated per group and analysed using logistic regression with study group as the main effect. The results will be presented as an odds ratio (95% CI).

#### 3.4.9 Secondary outcome: Overall quality of life

Measured at recruitment and PIVC removal by parent report, using the PedsQL™ Infant Scales (1–12 month version, 36 items) and/or the Infant Health-Related Quality of Life Instrument (IQI, 7 items); the IQI replaced the PedsQL™ from 23/05/2024. The PedsQL™ total score will be calculated per the scoring manual (items reverse-scored and transformed to 0–100, higher scores indicating better quality of life), excluding items for infants over 13 months. The IQI total score will be the sum of the seven item scores (range 7–28, higher scores indicating poorer health). Total scores will be summarised by group (mean (SD) or median (IQR)) at both time points, separately for each instrument, among participants who completed it. Between-group differences at PIVC removal will be estimated using linear regression adjusted for the baseline score and randomisation stratum, and presented as a mean difference (95% CI) for the IQI. As few participants completed the PedsQL™, its scores will be reported descriptively. Scores will be calculated when at least 50% of items are answered, per each instrument’s scoring guidance. Quality of life at 1, 3 and 6 months will be reported with the health economic evaluation.

#### 3.4.10 Safety outcome: Serious adverse events (including mortality, severe extravasation injury, and severe device-related injury)

The number and percentage of participants experiencing a serious adverse event will be reported per group and compared using Firth penalised logistic regression adjusted for randomisation stratum, presented as an odds ratio (95% CI), if at least one serious adverse event occurs; otherwise serious adverse events will be reported as counts. All serious adverse events will be reported narratively.

#### 3.4.11 Exploratory analysis: IV biosensor red alarm

Among participants in the IV biosensor group with an extravasation injury, extravasation severity (CCHMC) and volume will be compared between those with and without a red alarm before injury identification. Results will be summarised descriptively (n, %; median, IQR); if numbers permit, ordinal logistic regression (severity) and linear or quantile regression (volume) will be used. As this comparison is not randomised, it is hypothesis generating only. Alarms will be classed as preceding injury when an alarm time precedes the recorded time of injury identification; only the first three alarm times per daily check are recorded.

## 4. Protocol amendments and differences from the protocol

### 4.1 Protocol amendments

### 4.2 Differences between protocol v3.1 and this SAP

## 5. Trial status

The PATCH trial began recruitment on 01/11/2023. This original version of the SAP was finalised on 10/11/2026, before database lock and before any unblinded analysis of trial outcomes. Decisions informed by trial data used blinded, pooled data only. Recruitment was ongoing when this plan was finalised. The analysis of primary and secondary outcomes will be conducted upon completion of the trial. Prior to submission of the manuscript, the SAP will be updated to reflect any changes made during the analysis process. A list of changes between this original SAP and the final SAP will be detailed, along with the timing of, and reason for, the change.

## 6. Subsequent publications

Quality of life at 1, 3 and 6 months, extravasation-related quality of life and cost outcomes will be reported in a separate health economic evaluation, under its own analysis plan.

## Data Availability

Not applicable. This article is a statistical analysis plan and contains no data. Data from the trial will be reported in the primary results publication.

## 7. Declarations

### Ethics approval

Approved by the Children’s Health Queensland Human Research Ethics Committee (HREC/23/QCHQ/95263) and ratified by The University of Queensland (2023/HE001089). Parents or caregivers provide written informed consent.

## List of abbreviations

Abbreviation: Definition
ACF: antecubital fossa
ADLs: activities of daily living
ANZCTR: Australian New Zealand Clinical Trials Registry
CCHMC: Cincinnati Children’s Hospital Medical Center
CHQ: Children’s Health Queensland
CI: confidence interval
CONSORT: Consolidated Standards of Reporting Trials
CRF: case report form
DEM: department of emergency medicine
EBM: expressed breast milk
ED: emergency department
EQ-5D: EuroQol 5-Dimension
FLACC: Face, Legs, Activity, Cry, Consolability scale
GSNU: Grantley Stable Neonatal Unit
HR: hazard ratio
ICD-10: International Classification of Diseases, 10th revision
IQI: Infant Quality of Life Instrument
IQR: interquartile range
ITT: intention to treat
IV: intravenous
MGC: metric graduated colour
NGT: nasogastric tube
NICU: neonatal intensive care unit
OAC: Outcome Assessment Committee
OR: odds ratio
PATCH: Preventing InfusAte injuries Throughout a Child’s Hospitalisation
PedsQL: Pediatric Quality of Life Inventory
PICU: paediatric intensive care unit
PIPP: Premature Infant Pain Profile
PIVC: peripheral intravenous catheter
QAS: Queensland Ambulance Service
QCH: Queensland Children’s Hospital
QoL: quality of life
RBWH: Royal Brisbane and Women’s Hospital
RCT: randomised controlled trial
REDCap: Research Electronic Data Capture
RMO: resident medical officer
SAE: serious adverse event
SAP: statistical analysis plan
SCN: special care nursery
SCUH: Sunshine Coast University Hospital
SD: standard deviation

## Appendix 1. Proposed tables and figures

**Table 1.** Patient characteristics. Demographic and clinical data will be reported by the allocated study group.

| **Variables** |  | **Values** | **IV biosensor (N)** | **Standard Care (N)** |
|---|---|---|---|---|
| **Age at enrolment (days)** | **Mean (SD) or median (IQR)** |  |  |  |
| **Premature (<38 weeks)** | **n (%)** | Yes |  |  |
| **Gestational age at birth (weeks; infants born <38 weeks)** | **Mean (SD) or median (IQR)** |  |  |  |
| **Sex** | **n (%)** | Male |  |  |
|  |  | Female |  |  |
| **Aboriginal and/or Torres Strait Islander status** | **n (%); Multiple options** | Aboriginal Australian |  |  |
|  |  | Torres Strait Islander |  |  |
|  |  | Neither |  |  |
| **Weight (kg)** | **Mean (SD) or median (IQR)** |  |  |  |
| **Primary diagnosis category (ICD-10 chapter, where a discharge summary is available)** | **n (%)** | ICD-10 chapters |  |  |
| **Skin Colour using MGC tool** | **n (%)** | Peach |  |  |
|  |  | Blush |  |  |
|  |  | Pink |  |  |
|  |  | Yellow |  |  |
|  |  | Mustard |  |  |
|  |  | Orange |  |  |
|  |  | Fuchsia |  |  |
|  |  | Red |  |  |
|  |  | Tan |  |  |
|  |  | Brown |  |  |
|  |  | Chocolate |  |  |
|  |  | Violet |  |  |
|  |  | Purple |  |  |
|  |  | Black |  |  |

Reported in a supplementary table: breastfeeding status, cultural background, language spoken at home and country of birth. Unknown/prefer not to answer responses are treated as missing and reported in table footnotes. Categories with <5% of participants in both groups will be combined as 'Other'.

**Table 2.**
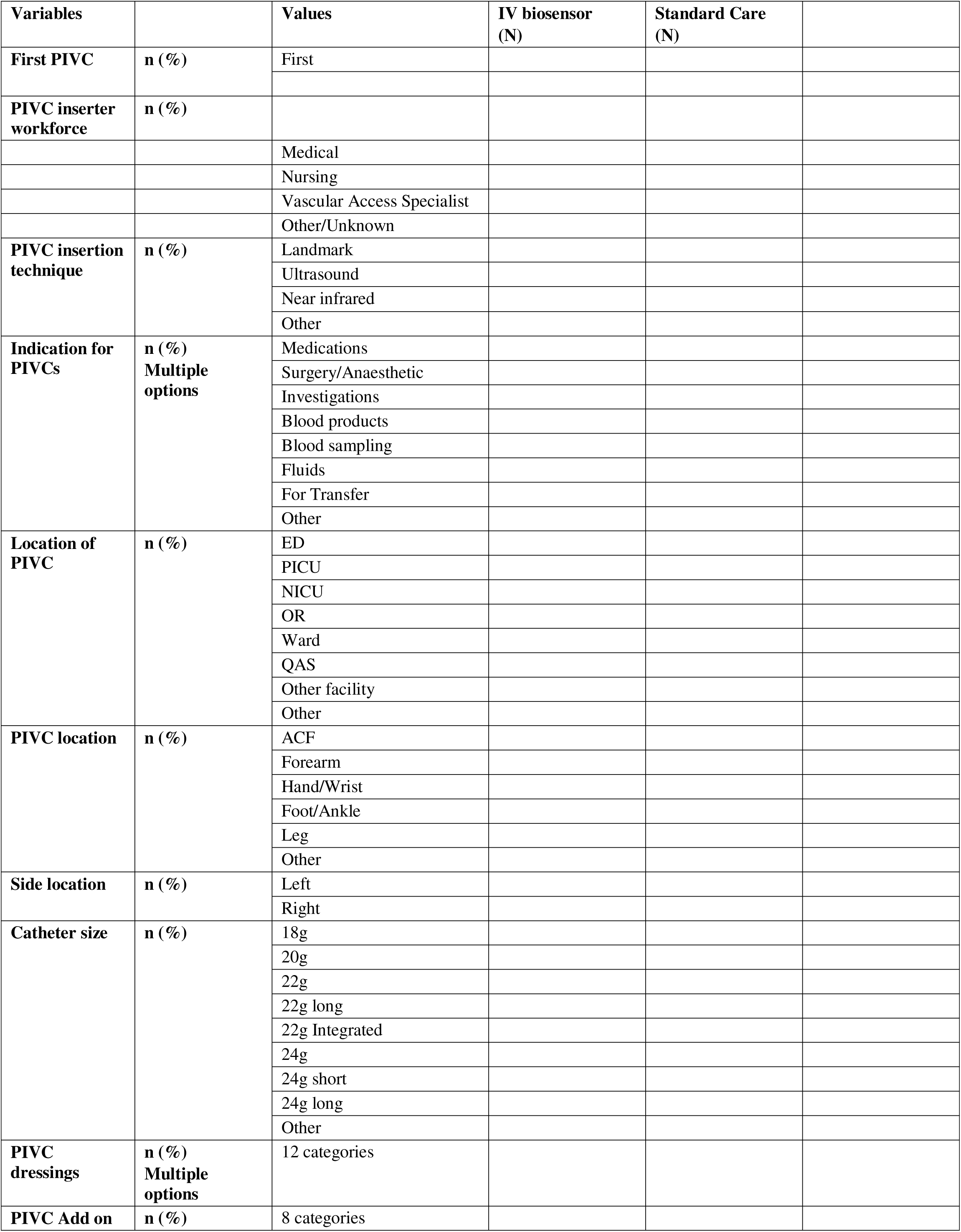
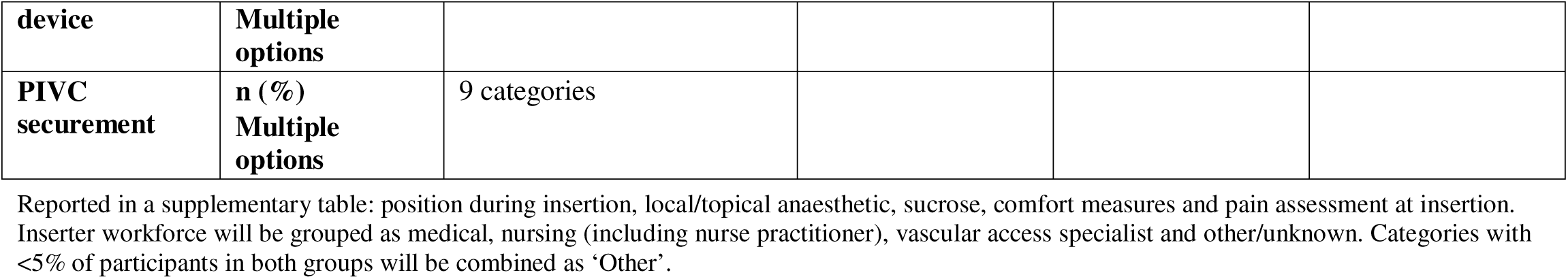
Device characteristics. Device characteristics collected at baseline will be reported by allocated study group.

| **Variables** |  | **Values** | **IV biosensor**<br>**(N)** | **Standard Care**<br>**(N)** |
|---|---|---|---|---|
| **First PIVC** | **n (%)** | First |  |  |
| **PIVC inserter workforce** | **n (%)** |  |  |  |
|  |  | Medical |  |  |
|  |  | Nursing |  |  |
|  |  | Vascular Access Specialist |  |  |
|  |  | Other/Unknown |  |  |
| **PIVC insertion technique** | **n (%)** | Landmark |  |  |
|  |  | Ultrasound |  |  |
|  |  | Near infrared |  |  |
|  |  | Other |  |  |
| **Indication for PIVCs** | **n (%)**<br>**Multiple options** | Medications |  |  |
|  |  | Surgery/Anaesthetic |  |  |
|  |  | Investigations |  |  |
|  |  | Blood products |  |  |
|  |  | Blood sampling |  |  |
|  |  | Fluids |  |  |
|  |  | For Transfer |  |  |
|  |  | Other |  |  |
| **Location of PIVC** | **n (%)** | ED |  |  |
|  |  | PICU |  |  |
|  |  | NICU |  |  |
|  |  | OR |  |  |
|  |  | Ward |  |  |
|  |  | QAS |  |  |
|  |  | Other facility |  |  |
|  |  | Other |  |  |
| **PIVC location** | **n (%)** | ACF |  |  |
|  |  | Forearm |  |  |
|  |  | Hand/Wrist |  |  |
|  |  | Foot/Ankle |  |  |
|  |  | Leg |  |  |
|  |  | Other |  |  |
| **Side location** | **n (%)** | Left |  |  |
|  |  | Right |  |  |
| **Catheter size** | **n (%)** | 18g |  |  |
|  |  | 20g |  |  |
|  |  | 22g |  |  |
|  |  | 22g long |  |  |
|  |  | 22g Integrated |  |  |
|  |  | 24g |  |  |
|  |  | 24g short |  |  |
|  |  | 24g long |  |  |
|  |  | Other |  |  |
| **PIVC dressings** | **n (%)**<br>**Multiple options** | 12 categories |  |  |
| **PIVC Add on device** | **n (%)**<br>**Multiple options** | 8 categories |  |  |
| **PIVC securement** | **n (%)**<br>**Multiple options** | 9 categories |  |  |

Reported in a supplementary table: position during insertion, local/topical anaesthetic, sucrose, comfort measures and pain assessment at insertion.
Inserter workforce will be grouped as medical, nursing (including nurse practitioner), vascular access specialist and other/unknown. Categories with <5% of participants in both groups will be combined as ‘Other’.

**Table 3.**
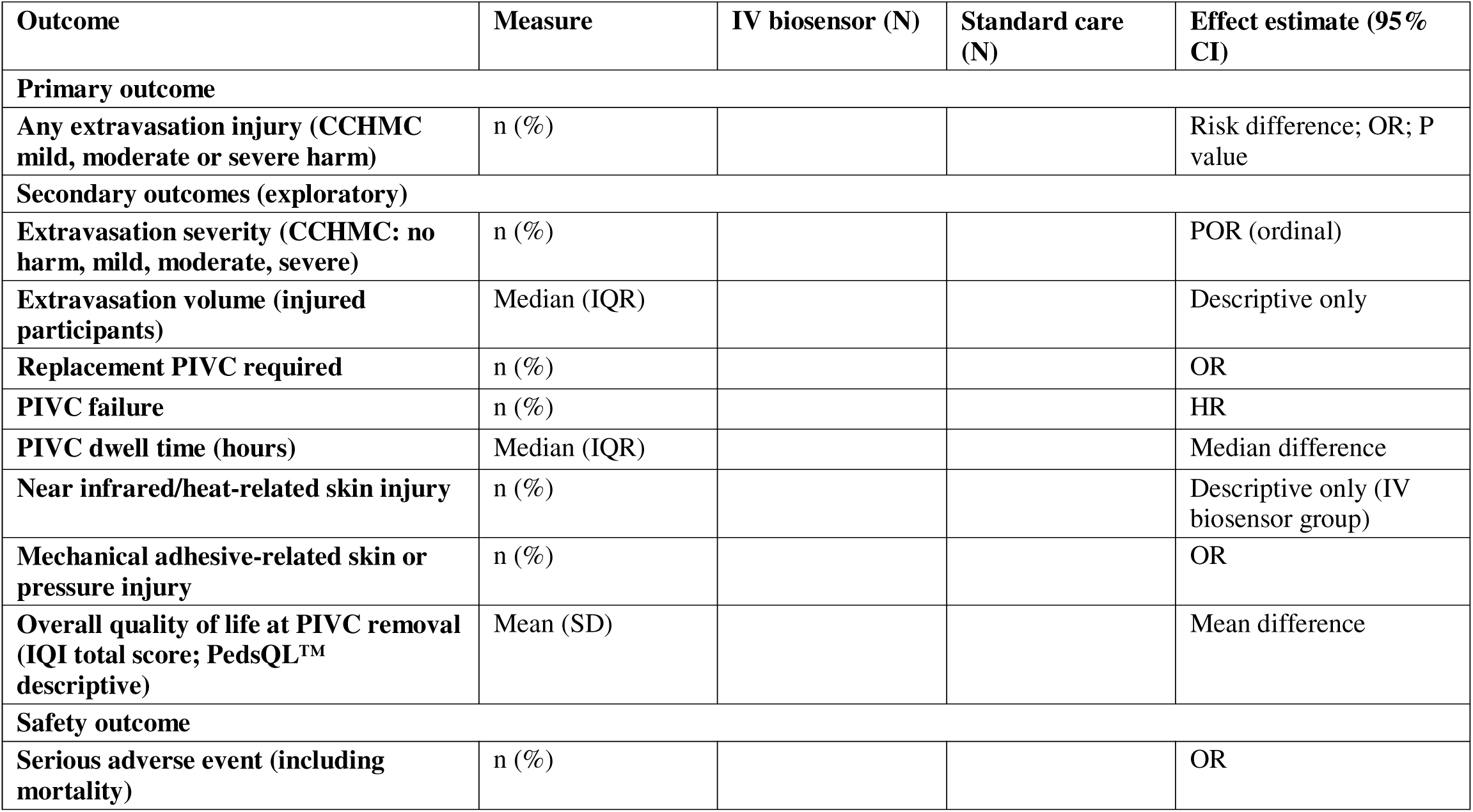
Primary, secondary and safety outcomes. Outcomes will be reported by allocated study group. All other tables will be supplementary. All models are adjusted for stratum; P values are reported for the primary outcome only. Cost outcomes and quality of life after PIVC removal are reported separately (health economic analysis plan). Extravasation treatment, pH and colour changes are measured only in injured participants and are reported descriptively in Table S3.

**Table 4.** Protocol amendments made during the trial.

| Version | Date | Changes relevant to the analysis |
|---|---|---|
| 1.0 | 16/04/2023 | Original protocol. Recruitment began 01/11/2023. |
| 2.0 | 12/02/2024 | Lipid-based infusate removed from the exclusion criteria. Biosensor placement changed from 10–15 mm of the PIVC tip to within 2.5 cm of the insertion site (4.5 cm for long PIVCs). Extravasation severity (four CCHMC categories) added as a secondary outcome. Quality of life instrument changed from EQ-5D to PedsQL™ Infant Scales (IQI from 23/05/2024), with 1, 3 and 6 month follow-up for injured participants only. Outcome Assessment Committee review added for suspected injuries, PIVC dysfunction and red alerts. pH sub-study added. |
| 2.1 | 13/09/2024 | Implementation interviews moved from the final year to during the trial. No effect on the effectiveness analysis. |
| 3.0 | 04/06/2025 | Minimum weight lowered from ≥1250 g to ≥1000 g. QCH recruitment widened beyond Ward 9B. |
| 3.1 | 28/04/2026 | Investigator added. No change to design or outcomes. |

CCHMC, Cincinnati Children’s Hospital Medical Center; EQ-5D, EuroQol 5-Dimension; IQI, Infant Quality of Life Instrument; PedsQL, Pediatric Quality of Life Inventory; PIVC, peripheral intravenous catheter; QCH, Queensland Children’s Hospital.

**Table 5.** Differences between protocol v3.1 and this SAP.

| **Item** | **Protocol v3.1** | **SAP** | **Rationale** |
|---|---|---|---|
| Primary effect measure | Logistic regression (OR) | Risk difference by standardisation after logistic regression; OR also reported | Absolute effect is easier to interpret clinically |
| QoL and cost outcomes | Listed as secondary outcomes | Overall QoL at recruitment and PIVC removal analysed here; QoL at 1, 3 and 6 months, BBSIP and cost in a separate health economic analysis plan | Reported in a subsequent publication |
| PIVC dwell time | Mean dwell time | Time to PIVC failure (HR) and median difference | Dwell time is skewed; failure is censored |
| Extravasation volume, treatment, pH and colour | Between-group comparison | Descriptive only (Table S3) | Measured only in injured participants |
| Replacement PIVC | Participants with PIVC removed for dysfunction | All participants (yes/no) | Avoids conditioning on a post-randomisation event |
| Near infrared skin injury | Logistic regression (OR) | Descriptive, IV biosensor group only | Injury can occur only with the sensor; REDCap field not displayed |
| Red alarm analysis | Not specified | Exploratory, IV biosensor group with injury | Hypothesis generating |

HR, hazard ratio; OR, odds ratio; PIVC, peripheral intravenous catheter; QoL, quality of life; SAP, statistical analysis plan.

## Supplementary tables

**Table S1.** Daily observation characteristics. Daily observation characteristics throughout dwell will be reported by allocated study group. These are post-randomisation process measures and will be reported descriptively, without between-group comparison. IV biosensor alarms (IV biosensor group only) will be reported in a separate supplementary table.

| Variables |  | **Values** | **IV biosensor (N)** | **Standard Care (N)** |
|---|---|---|---|---|
| **Use of PIVC** | **n (%)** | Fluids |  |  |
|  |  | Medications |  |  |
|  |  | Blood products |  |  |
|  |  | Blood sampling |  |  |
|  |  | Bunged |  |  |
|  |  | Investigation |  |  |
|  |  | Others |  |  |
| **Number of times the PIVC was accessed (Not including flushes)** | **Mean (SD) or median (IQR)** |  |  |  |
| **Infusion therapy used majority of time** | **n (%)** | Continuous |  |  |
|  |  | Intermittent |  |  |
|  |  | Continuous with intermediate medications |  |  |
|  |  | None |  |  |
| **Maximum rate IVF (ml/hr)** | **Mean (SD) or median (IQR)** |  |  |  |
| **Medications received** | **n (%)<br>Multiple options** | 25+ categories |  |  |
| **Antimicrobials** | **n (%)<br>Multiple options** | 25+ categories |  |  |
| **Infusates** | **n (%)** | No (low risk only - all green) |  |  |
|  |  | Yes (intermediate risk - any yellow) |  |  |
|  |  | Yes (high risk - any red) |  |  |
|  |  | Unknown |  |  |
| **Dressing reinforcement** | **n (%)** | Yes |  |  |
|  |  | No |  |  |
| **IV disconnection** | **n (%)** | Yes |  |  |
|  |  | No |  |  |
|  |  | Unknown |  |  |

**Table S2.** Removal characteristics. Removal characteristics at the end of the study will be reported by allocated study group. Outcomes are reported in Table 3.

| **Variables** |  | **Values** | **IV biosensor<br>(N)** | **Standard Care<br>(N)** |
|---|---|---|---|---|
| **Dwell (days)** | **Mean (SD) or median (IQR)** |  |  |  |
| **Reason for removal** | **n (%)<br>Multiple options** | Completed therapy |  |  |
|  |  | Extravasation/ infiltration |  |  |
|  |  | Non-extravasation related PIVC dysfunction |  |  |
|  |  | Palliation |  |  |
|  |  | Death |  |  |
|  |  | Other |  |  |
|  |  | Unknown |  |  |
| **Number of PIVCs required** | **n (%)** | 1 |  |  |
|  |  | 2 |  |  |
|  |  | 3 |  |  |
|  |  | 4 |  |  |
|  |  | 5 |  |  |
|  |  | Unknown |  |  |
| **Type of skin injury** | **n (%)** | Skin tear |  |  |
|  |  | Stage 1 Pressure Injury |  |  |
|  |  | Stage 2 Pressure injury |  |  |
|  |  | Other |  |  |
| **SAE type** | **n (%)** | Mortality |  |  |
|  |  | Severe extravasation injury (grade 3 or higher) |  |  |
|  |  | Severe device related injury – cord injury |  |  |
|  |  | Severe device related injury -near infra-red burn |  |  |
|  |  | Other |  |  |

**Table S3.** Extravasation characteristics. Extravasation characteristics for cases will be reported by allocated study group. Participants with an injury only.

| **Variables** |  | **Values** | **IV biosensor**<br>**(N)** | **Standard Care**<br>**(N)** |
|---|---|---|---|---|
| **Methods of extravasation identification** | **n (%)** | Clinical assessment |  |  |
|  |  | IV watch, confirmed by clinical assessments |  |  |
| **Paediatric PIV infiltration scale** | **n (%)** | Grade 0 |  |  |
|  |  | Grade 1 |  |  |
|  |  | Grade 2 |  |  |
|  |  | Grade 3 |  |  |
|  |  | Grade 4 |  |  |
| **Extravasation volume (ml)** | **Mean (SD) or median (IQR)** |  |  |  |
| **pH at injury site** | **Mean (SD) or median (IQR)** |  |  |  |
| **Skin Colour using MGC tool** | **n (%)** | Peach |  |  |
|  |  | Blush |  |  |
|  |  | Pink |  |  |
|  |  | Yellow |  |  |
|  |  | Mustard |  |  |
|  |  | Orange |  |  |
|  |  | Fuchsia |  |  |
|  |  | Red |  |  |
|  |  | Tan |  |  |
|  |  | Brown |  |  |
|  |  | Chocolate |  |  |
|  |  | Violet |  |  |
|  |  | Purple |  |  |
|  |  | Black |  |  |
| **Length of stay (days)** | **Mean (SD) or median (IQR)** |  |  |  |
| **Extravasation treatment** | **n (%)** |  |  |  |
| **Dressing interventions required** | **n (%)** |  |  |  |
| **Medical interventions required** | **n (%)** |  |  |  |
| **Surgical interventions required** | **n (%)** |  |  |  |
| **Analgesia required** | **n (%)** |  |  |  |
| **Red alarm before injury identification** | **n (%)** |  |  |  |

SAP PATCH v.original

---

**Figure A1.**
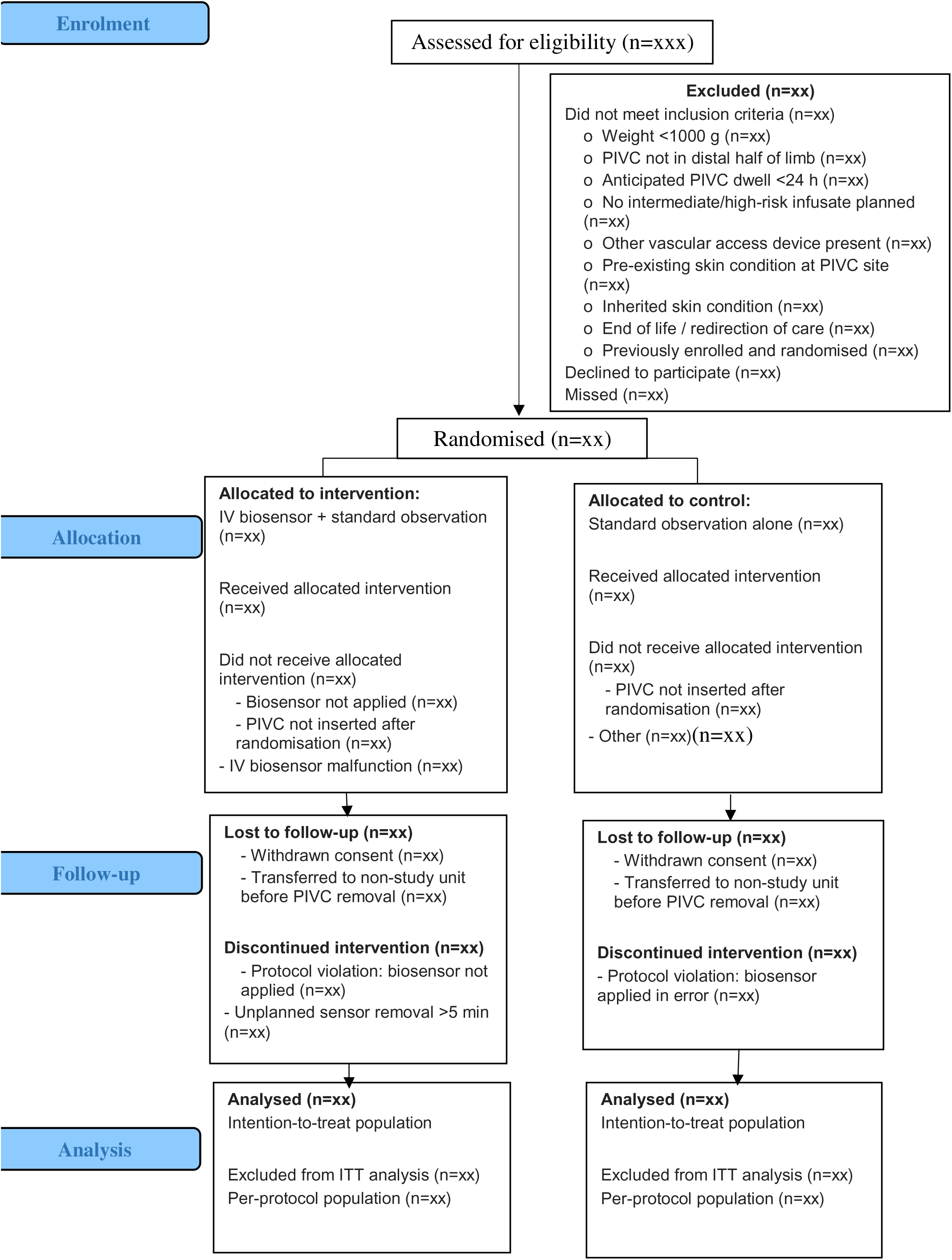
CONSORT participant flow diagram

## Appendix 2. Data dictionary

Variable definitions, categories, data types and data sources for all variables reported in this SAP

### A2.1 Patient characteristics

| **Variable** | **Description** | **Data type** | **Method of presentation** | **Variable name** | **Instrument** |
|---|---|---|---|---|---|
| Site | QCH, RBWH, SCUH | Nominal | Counts and percentages | ed_site | Enrolment |
| Ward/unit (SCUH only) | SCUH NU/SCN, SCUH Paeds | Nominal | Counts and percentages | ed_dem_ward_location | Enrolment |
| Randomisation stratum | QCH, RBWH, SCUH Neonatal Unit, SCUH Children’s and Adolescent Unit | Nominal | Counts and percentages | Derived from ed_site and ed_dem_ward_location | Enrolment |
| Allocated group | IV biosensor, Standard care | Nominal | Counts and percentages | ed_allocated_group | Enrolment |
| Date of birth | Date | — | Used to derive age | ed_dem_dob | Enrolment |
| Age at enrolment (days) | Days | Continuous | Median (IQR) and range | Derived from ed_dem_dob | Enrolment |
| Premature | Yes, No | Nominal | Counts and percentages | ed_dem_prem | Enrolment |
| Estimated due date | Date | — | Used to derive corrected age | ed_edd | Enrolment |
| Sex | Male, Female, Other | Nominal | Counts and percentages | ed_dem_gender | Enrolment |
| Weight (kg) | kg | Continuous | Mean (SD) or median (IQR) | ed_dem_wt_kg | Enrolment |
| Breastfeeding status | Exclusive breastfeeding (including EBM via bottle or NGT), Some breastfeeding, Not breastfeeding | Nominal | Counts and percentages | ed_dem_bf | Enrolment |
| Aboriginal and/or Torres Strait Islander status | Aboriginal, Torres Strait Islander, Both, Neither | Nominal | Counts and percentages | ed_dem_atsi_status | Enrolment |
| Cultural background | Multiple categories | Nominal | Counts and percentages | pd_cultural_background | Enrolment |
| Language spoken at home | Multiple categories | Nominal | Counts and percentages | ed_dem_language | Enrolment |
| Country of birth | Australia, NZ and Pacific Islands, UK and Republic of Ireland, Europe, Asia, Americas, Africa and Middle East, Unknown/prefer not to answer | Nominal | Counts and percentages | ed_dem_country_of_birth | Enrolment |
| Skin colour (MGC tool) | Peach, Blush, Pink, Yellow, Mustard, Orange, Fuchsia, Red, Tan, Brown, Chocolate, Violet, Purple, Black, White | Nominal | Counts and percentages | ed_dem_skincolour | Enrolment |

### A2.2 Device characteristics

| **Variable** | **Description** | **Data type** | **Method of presentation** | **Variable name** | **Instrument** |
|---|---|---|---|---|---|
| Date of PIVC insertion | Date | — | Used to derive dwell time | ed_iv_dateinsertion | Enrolment |
| First or subsequent PIVC | First, Subsequent | Nominal | Counts and percentages | ed_iv_picvnumber | Enrolment |
| Indication/planned utility | Medications, Surgery/Anaesthetic, Investigations, Blood products, Blood sampling, Fluids, For transfer, Other (multiple) | Nominal | Counts and percentages | ed_iv_utility | Enrolment |
| Inserter workforce | Treating team RMO/intern, Registrar/fellow, Consultant, ACE registrar, Anaesthetics doctor, DEM doctor, DEM nurse, PICU doctor, NICU doctor, Vascular Access Specialist, Nurse, Nurse Practitioner, Radiographer, QAS, Other, Unknown | Nominal | Counts and percentages | ed_iv_inserter_workforce | Enrolment |
| Insertion technique | Landmark, Ultrasound, Near Infrared, Transilluminator, Other, Unknown | Nominal | Counts and percentages | ed_iv_insertiontech | Enrolment |
| Number of insertion attempts | 1, 2, 3, 4, 5, >5, Unknown | Ordinal | Counts and percentages | ed_iv_attempts | Enrolment |
| Clinical area of insertion | ED, PICU, NICU/SCN, OR, Ward, QAS, Delivery suite, Other facility, Other | Nominal | Counts and percentages | ed_iv_insert_location | Enrolment |
| Documentation of insertion | Documented in notes, Parent/carer report, Both, Neither | Nominal | Counts and percentages | ed_iv_document | Enrolment |
| PIVC anatomical location | ACF, Forearm, Hand, Wrist, Foot, Ankle, Lower leg | Nominal | Counts and percentages | ed_iv_location | Enrolment |
| Side of insertion | Left, Right | Nominal | Counts and percentages | ed_iv_side_location | Enrolment |
| Catheter size | 18g, 20g, 22g standard, 22g Long, 22g Integrated, 24g standard, 24g Long, 26g, Other | Nominal | Counts and percentages | ed_iv_catheter | Enrolment |
| Arm length at PIVC site (cm) | cm | Continuous | Mean (SD) or median (IQR) | ed_injury_arm_length_y | Enrolment |
| PIVC dressings | Multiple categories | Nominal | Counts and percentages | ed_iv_dressings | Enrolment |
| Add-on devices | Multiple categories | Nominal | Counts and percentages | ed_iv_adddevices | Enrolment |
| Green spot applied | Yes, No | Nominal | Counts and percentages | ed_iv_greenspot | Enrolment |

### A2.3 Provider characteristics

| Variable | Description | Data type | Method of presentation | Variable name | Instrument |
|---|---|---|---|---|---|
| Pain assessment performed at insertion | Yes, No, Unknown, Unable to assess | Nominal | Counts and percentages | ed_pain_assessment | Enrolment |
| Pain assessment tool used | FLACC, PIPP, NAIPAAS, MPAT, Other (multiple) | Nominal | Counts and percentages | ed_pain_assessment_tool | Enrolment |
| FLACC score | Numeric | Continuous | Median (IQR) | pain_score_flacc | Enrolment |
| PIPP score | Numeric | Continuous | Median (IQR) | pain_score_pipp | Enrolment |
| NAIPAAS score | Numeric | Continuous | Median (IQR) | pain_score_naipas | Enrolment |
| MPAT score | Numeric | Continuous | Median (IQR) | pain_score_mpat | Enrolment |
| Local anaesthetic used | Yes, No, Unknown | Nominal | Counts and percentages | ed_pain_anaesthetics | Enrolment |
| Local anaesthetic type | Multiple categories | Nominal | Counts and percentages | ed_pain_anaesthetics_list | Enrolment |
| Sucrose used | Yes, No, Unknown | Nominal | Counts and percentages | ed_pain_sucrose | Enrolment |
| Comfort measures used | Yes, No, Unknown | Nominal | Counts and percentages | ed_pain_comfort | Enrolment |
| Type of comfort measures | Skin to skin, Breastfeeding, Swaddling, Pacifier, Other (multiple) | Nominal | Counts and percentages | ed_pain_comfort_list | Enrolment |
| Position during insertion | Held by parents, On bed, Unknown | Nominal | Counts and percentages | ed_pain_position | Enrolment |

### A2.4 Daily observation characteristics

Daily observation data are collected each day the PIVC is in situ and will be reported summarised across the dwell period by allocated study group.

| Variable | Description | Data type | Method of presentation | Variable name | Instrument |
|---|---|---|---|---|---|
| PIVC daily utility | Fluids, Medications, Blood products, Blood sampling, Bunged, Investigation, Other (multiple) | Nominal | Counts and percentages | dc_daily_utility | Daily check |
| PIVC access frequency | 0, 1–5, 6–10, >10, Unknown | Ordinal | Counts and percentages | dc_pivc_accessed | Daily check |
| Infusate risk level | Green (low risk), Yellow (intermediate risk), Red (high risk) | Nominal | Counts and percentages | dc_traffic_light | Daily check |
| Infusate therapy type | Continuous, Intermittent, Continuous with intermediate medications, None/saline flushes only | Nominal | Counts and percentages | dc_infusate_therapy | Daily check |
| Maximum infusate rate (mL/hr) | mL/hr | Continuous | Mean (SD) or median (IQR) | dc_infusate_rate | Daily check |
| PIVC disconnection occurred | Yes, No, Unknown | Nominal | Counts and percentages | dc_disconnections | Daily check |
| Dressing or device change | Yes, No | Nominal | Counts and percentages | dc_changesdevice | Daily check |
| PIVC dysfunction identified | Yes, No | Nominal | Counts and percentages | dc_piv_dysfunction | Daily check |
| Type of PIVC dysfunction | Multiple categories (infiltration/extravasation, occlusion, dislodgement, phlebitis, leaking, etc.) | Nominal | Counts and percentages | dc_dysfunction_type | Daily check |
| Protocol violation | Yes, No | Nominal | Counts and percentages | dc_protocol_violation | Daily check |
| IV biosensor removed during dwell | No; Yes <5 min remain off; Yes <5 min back in situ; Yes >5 min remain off; Yes >5 min back in situ; Unknown | Nominal | Counts and percentages | bw_ivwatch_removed; dc_ivwatch_removed | Daily check |
| Reason for IV biosensor removal | Alarmed, Device malfunction, ADLs (including bath), Other | Nominal | Counts and percentages | dc_ivwatch_removed | Daily check |
| IV biosensor probe in situ | Yes, No | Nominal | Counts and percentages | bw_ivwatch_probe | Daily check |
| IV biosensor alarm occurred | Yes, No | Nominal | Counts and percentages | dc_alarms | Daily check |
| Total daily alarms | Count | Count | Median (IQR) | dc_totaldailyalarms | Daily check |
| Alarm type | Yellow, Red, Other | Nominal | Counts and percentages | dc_alarm1_type; dc_alarm2_type; dc_alarm3_type | Daily check |
| Alarm non-clinical | Yes, No, Unknown | Nominal | Counts and percentages | dc_alarm1_nonclinical; dc_alarm2_nonclinical; dc_alarm3_nonclinical | Daily check |
| Non-clinical alarm cause | Battery, Sensor, Cable, Excessive ambient light, Other | Nominal | Counts and percentages | dc_nonclinical_alarm1–3 | Daily check |

### A2.5 Removal characteristics

Removal characteristics collected at PIVC removal will be reported by allocated study group.

| Variable | Description | Data type | Method of presentation | Variable name | Instrument |
|---|---|---|---|---|---|
| Reason for study completion | IV removed, Withdrawn by participant, Clinical decision to withdraw, Transferred to other facility, Other | Nominal | Counts and percentages | ivoc_reason_completion | PIVC outcomes 24h |
| Reason for PIVC removal | Completed therapy, Infiltration/extravasation, Non-infiltration/extravasation dysfunction, Palliation, Death, Other, Unknown (multiple) | Nominal | Counts and percentages | ivoc_removed_reason_end | PIVC outcomes 24h |
| PIVC dwell time (hours) | Hours | Continuous | Median (IQR) | Derived from ivoc_dtremovediv and ed_iv_dateinsertion | PIVC outcomes 24h |
| Replacement PIVC required | Yes successfully resited, Yes but unsuccessful, No not required | Nominal | Counts and percentages | ivoc_new_ac_end | PIVC outcomes 24h |
| Near infrared/heat related skin injury | Yes, No | Nominal | Counts and percentages | ivoc_infaredinjury | PIVC outcomes 24h |
| Pressure injury | Yes, No | Nominal | Counts and percentages | ivoc_pi | PIVC outcomes 24h |
| Type of skin injury | Multiple categories (skin tear, Stage 1 pressure injury, Stage 2 pressure injury, other) | Nominal | Counts and percentages | ivoc_skininjury | PIVC outcomes 24h |
| Serious adverse event occurred | Yes, No | Nominal | Counts and percentages | ivoc_sae_yes | PIVC outcomes 24h |
| Serious adverse event type | Mortality, Extravasation injury (amber or higher), Severe device-related injury – cord injury, Severe device-related injury – near infrared burn | Nominal | Counts and percentages | ivoc_sae | PIVC outcomes 24h |

### A2.6 Infiltration/extravasation injury characteristics

Reported for participants with a confirmed injury only, by allocated study group.

| Variable | Description | Data type | Method of presentation | Variable name | Instrument |
|---|---|---|---|---|---|
| Method of injury identification | Clinical assessment; ivWatch confirmed by clinical assessment | Nominal | Counts and percentages | ivoc_ex_end | PIVC outcomes 24h |
| PIVC in situ at time of injury identification | Yes, No, Unknown | Nominal | Counts and percentages | ivoc_insitu | PIVC outcomes 24h |
| Injury length (cm) | cm | Continuous | Median (IQR) | ivoc_injurylength_x | PIVC outcomes 24h |
| Arm length at injury site (cm) | cm | Continuous | Median (IQR) | ivoc_injury_arm_length_y | PIVC outcomes 24h |
| Extravasation volume (mL) | mL | Continuous | Median (IQR) | ivoc_extravasation_vol | PIVC outcomes 24h |
| Skin colour at injury site (MGC tool) | Peach, Blush, Pink, Yellow, Mustard, Orange, Red, Tan, Brown, Chocolate, Violet, Purple, Black, White, Fuchsia | Nominal | Counts and percentages | ivoc_ex_skincolour | PIVC outcomes 24h |
| pH at injury site | Numeric | Continuous | Median (IQR) | ivoc_ex_ph | PIVC outcomes 24h |
| Limb impairment | Yes, No | Nominal | Counts and percentages | ivoc_limb_impairment | PIVC outcomes 24h |
| Arterial compromise | Yes, No | Nominal | Counts and percentages | ivoc_artcompro | PIVC outcomes 24h |
| Limb colour (affected) | Pink, Pale, Flushed, Dusky, Mottled, Cyanotic, Grey, Jaundice, Normal for race, Unable to assess | Nominal | Counts and percentages | ivoc_limb_col | PIVC outcomes 24h |
| Limb colour (unaffected comparator) | As above | Nominal | Counts and percentages | ivoc_limb_col2 | PIVC outcomes 24h |
| Limb temperature | Hot, Warm, Cool, Cold, Clammy, Diaphoretic, Unable to assess | Nominal | Counts and percentages | ivoc_limb_temp | PIVC outcomes 24h |
| Capillary refill time | <2 sec, 2–4 sec, 4–6 sec, 6–8 sec, >8 sec | Ordinal | Counts and percentages | ivoc_cap_ref | PIVC outcomes 24h |
| Limb swelling | Full, Hollow or wrinkled, Distended or hard | Ordinal | Counts and percentages | ivoc_limb_swell | PIVC outcomes 24h |
| Limb pulse | Present, Absent, N/A | Nominal | Counts and percentages | ivoc_limb_pulse | PIVC outcomes 24h |
| CCHMC Extravasation Harm Scale — mild harm criteria | Multiple checkboxes | Nominal | Counts and percentages | ivoc_mild_harm_end | PIVC outcomes 24h |
| CCHMC Extravasation Harm Scale — moderate harm criteria | Multiple checkboxes | Nominal | Counts and percentages | ivoc_moderate_harm_end | PIVC outcomes 24h |
| CCHMC Extravasation Harm Scale — severe harm criteria | Multiple checkboxes | Nominal | Counts and percentages | ivoc_severe_harm_end | PIVC outcomes 24h |
| Pediatric PIV Infiltration Scale grade | No harm, Mild harm (green), Moderate harm (amber), Severe harm (red) | Ordinal | Counts and percentages | po_piv_infil_scale (Committee); ivoc_ped_piv_infiltration_scale_end (research nurse) | PIVC outcomes 24h |
| Injury SAE flag | Yes, No | Nominal | Counts and percentages | sir_inj_sae | Suspected injury report |

### A2.7 Quality of life

Collected at recruitment and PIVC removal; reported by allocated study group, separately for each instrument.

| **Variable** | **Description** | **Data type** | **Method of presentation** | **Variable name** | **Instrument** |
|---|---|---|---|---|---|
| PedsQL™ Infant Scales total score | 36 items (1–12 month version), 0–4 response; transformed to 0–100 | Continuous | Mean (SD) or median (IQR) | ed_pql_*;<br>ivoc_pql_* | Enrolment; PIVC outcomes 24h |
| IQI total score | 7 items (sleeping, feeding, breathing, stooling, mood, skin, interaction), 1–4 response; sum 7–28 | Continuous | Mean (SD) or median (IQR) | ed_iqi_*;<br>eq_iqi_*;<br>ivoc_iqi_* | Enrolment; PIVC outcomes 24h |

